# Mental Health in UK Biobank Revised – development, implementation and results from an online questionnaire completed by 157,366 participants

**DOI:** 10.1101/19001214

**Authors:** Katrina A. S. Davis, Jonathan R. I. Coleman, Mark Adams, Naomi Allen, Gerome Breen, Breda Cullen, Chris Dickens, Elaine Fox, Nick Graham, Jo Holliday, Louise M Howard, Ann John, William Lee, Rose McCabe, Andrew McIntosh, Robert Pearsall, Daniel J. Smith, Cathie Sudlow, Joey Ward, Stan Zammit, Matthew Hotopf

## Abstract

This paper corrects and updates a paper published in BJPsych Open 2018 “Mental Health in UK Biobank” (https://doi.org/10.1192/bjo.2018.12) that was voluntarily retracted following the finding of errors in the coding of the variable for alcohol use disorder. Notably, the percentage of participants reaching threshold for alcohol use disorder on the Alcohol Use Disorder Identification Tool increased from 7% to 21%.

**Background:** UK Biobank is a well-characterised cohort of over 500,000 participants that offers unique opportunities to investigate multiple diseases and risk factors. An online mental health questionnaire completed by UK Biobank participants expands the potential for research into mental disorders.

**Methods:** An expert working group designed the questionnaire, using established measures where possible, and consulting with a service user group regarding acceptability. Operational criteria were agreed for defining likely disorder and risk states, including lifetime depression, mania/hypomania, generalised anxiety disorder, unusual experiences and self-harm, and current post-traumatic stress and alcohol use disorders.

**Results:** 157,366 completed online questionnaires were available by August 2017. Comparison of self-reported diagnosed mental disorder with a contemporary study shows a similar prevalence, despite respondents being of higher average socioeconomic status. Lifetime depression was the most common finding in 24% of participants (37,434), with current alcohol use disorder criteria met by 21% (32,602), while other criteria were met by less than 8% of the participants. There was extensive comorbidity among the syndromes. Mental disorders were associated with a high neuroticism score, adverse life events and long-term illness; addiction and bipolar affective disorder in particular were associated with measures of deprivation.

**Conclusions:** The questionnaire represents a very large mental health survey in itself, and the results presented here show high face validity, although caution is needed due to selection bias. Built into UK Biobank, these data intersect with other health data to offer unparalleled potential for crosscutting biomedical research involving mental health.

## Introduction

UK (United Kingdom) Biobank is a very large, population-based cohort study established to identify the determinants of common life-threatening and disabling conditions.^1^ Most of these conditions, like heart disease, stroke and mental disorders, are multifactorial, involving multiple genes of small effect, and complex relationships with environmental exposures. This means large samples are required to study associations between these exposures and disease, and identify targets for treatment and prevention.^2^ The utility of traditional epidemiological study designs is often limited by their focus on single disorders or exposures and relatively modest sample sizes.^3^ UK Biobank is an open-access resource providing detailed characterisation of over half a million people aged 40-69 years at recruitment, with proposed long term follow-up. Recruitment was completed in 2010, along with consent for future contact and linkage to routinely collected health-related data, such as those produced by the National Health Service (NHS). Baseline measures were extensive, from family history to sensory acuity (a searchable breakdown at www.ukbiobank.ac.uk), and the resource continues to grow. In 2017, genotyping of the whole cohort was complete, a range of blood biomarkers were released in 2019, and multimodal imaging is underway for 100,000 participants. Locality environmental factors, such as air pollution, are also available. The design of UK Biobank offers the opportunity to examine a wide range of risk factors and outcomes in a sample that has the size to provide the power to detect small effects, making UK Biobank a highly efficient resource for observational epidemiology.

The impact of mental disorders on disability and quality of life is considerable,^4^ accounting for the equivalent of over 1.2 million person years lost to disability from mental and substance use disorders in England alone in 2013^5^. The potential detrimental impact of mental disorders both on physical disease onset and outcomes^6-8^ is particularly notable for this project. The availability of mental health phenotypes in conjunction with the wealth of other data in the UK Biobank would offer considerable opportunities to study aetiological and prognostic factors. The UK Biobank baseline data collection of mental health, consisted of several questions about mood and a neuroticism scale, expanded for the last 172,729 recruited participants with questions to allow provisional categorisation of mood disorder;^9^ however, there was considerable scope for further characterisation of mental disorders among participants.

Characterising mental disorders in a cohort such as UK Biobank poses challenges. Firstly, most mental disorders manifest before age 30 years and have fluctuating courses,^10^ so a “snapshot” of disorder status at one point in time, as identified by most screening tools, is likely to be less useful than a “lifetime” history. Secondly, traditional diagnostic approaches to mental disorders, relying upon clinician assessment at interview, would be prohibitively expensive in a cohort of this size. Thirdly, using self-report of diagnosis or data from record linkages relies upon recognition of illness and reflects healthcare usage patterns, whereas many people with mental disorders never seek or receive treatment.^10, 11^ In response to these challenges, we developed a dual approach: secondary care record linkage for identification of more severe illnesses such as schizophrenia^12^ and self-report of symptoms of common mental disorder, which might not have come to clinical attention. As part of our mental health phenotyping programme we therefore developed an online mental health questionnaire (MHQ) for participants to complete regarding lifetime symptoms of mental disorders. The MHQ aimed to exploit the efficiency of “e-surveys”^13^ and provide the detail needed to identify mental health disorders without the need for a clinical assessment.

The present paper aims to describe the development, implementation and results of this questionnaire. We provide descriptive data on the numbers of UK Biobank participants meeting diagnostic criteria for specific disorders, and on the frequency of exposure to risk factors. We also evaluate the likely representativeness of respondents by comparing respondent socio-demographic characteristics to that of the UK population using census data, and comparing self-reported mental disorder diagnosis with the Health Survey for England (HSE) data.^14^ This will assist researchers considering or undertaking epidemiological research to evaluate the potential and power of using UK Biobank to look at mental health.

## Methods

### Questionnaire development

A mental health research reference group formed of approximately 50 individuals (see supplementary material Appendix 1) participated in discussions about a strategy for mental health phenotyping in UK Biobank, including a workshop in January 2015. From this, a smaller steering group was established and led the development of the mental health questionnaire (MHQ). The group recommended that the MHQ should concentrate on depression, as it was likely to represent the greatest burden in the cohort, with some questions about other common disorders, including anxiety, alcohol misuse and addiction, plus risk factors for mental disorder not captured at participants’ baseline assessment.

The intention was to create a composite questionnaire out of previously existing and validated measures, taking into account participant acceptability (time, ease of use, and ensuring questions were unlikely to offend), scope for collaborations with international studies (e.g. the Psychiatric Genomics Consortium) through making results comparable, and the need to balance depth and breadth of phenotyping. The base of the questionnaire was the measurement of lifetime depressive disorder using the Composite International Diagnostic Interview Short Form (CIDI-SF),^15^ modified to provide lifetime history, as used to identify cases and controls for some existing studies in the Psychiatric Genomics Consortium. The CIDI-SF uses a branching structure with screening questions and skip rules to limit detailed questions to the relevant areas for each participant. Other measures were then added to this, as summarised in supplementary material Table SM1. Where the group were unable to find existing measures that fulfilled these criteria, questions were written or adapted, as indicated in SM1. These sections have not been externally validated, but the questions can be seen, along with the full questionnaire on the UK Biobank website (http://biobank.ctsu.ox.ac.uk/crystal/refer.cgi?id=22), for researchers to evaluate how they wish to use them.

### Testing and ethical approval

The use of branching questions in the MHQ means that those with established and multiple mental disorders have a longer, more detailed, questionnaire. To improve acceptability in this group, we worked with a service user advisory group at the National Institute of Health Research (NIHR) Biomedical Research Centre at the South London and Maudsley (SLaM) NHS Foundation Trust in designing the questionnaire and invitation.^16^ We then piloted the questionnaire amongst an online cohort of 14,836 volunteers aged over 50 and living in the UK, who completed the questionnaire as part of signing up to take part in the Platform for Research Online to investigate Genetics and Cognition in Ageing (PROTECT).^17^ Of those who started the questionnaire 98.8% completed it, taking a median time of 15 minutes. Some PROTECT participants commented that they wanted the opportunity to explain why they felt they had experienced symptoms of depression. In response to this, we added a question to the depression section on loss or bereavement, and a free-text box – neither were designed to change diagnostic algorithms, but may add to future analyses.

The questionnaire was approved as a substantial amendment to UK Biobank approval from the North West - Haydock Research Ethics Committee, 11/NW/0382.

### Administration to UK Biobank Participants

We incorporated the final MHQ into the UK Biobank web questionnaire platform and presented it to participants as an online questionnaire entitled “thoughts and feelings”. We sent participants who had agreed to email contact a hyperlink to their personalised questionnaire. The invitation explained the importance of collecting further information about mental health and emphasised that UK Biobank was unable to respond to concerns raised by the participant in the questionnaire, instead directing them to several sources of potential support. Participants could skip questions they preferred not to answer, and they could save answers to return to the questionnaire later. We sent reminder emails at two weeks and four months to those who had not started or had partially completed the questionnaire. The MHQ will continue to be available on the participant area of the UK Biobank website, and since 2017 the annual postal newsletter contains an invitation to log on to the participant area and complete questionnaires, which will reach those for whom no email contact was possible. Data from the MHQ will therefore continue to accrue. The current numbers and aggregate data can be accessed from the public data showcase (http://biobank.ctsu.ox.ac.uk/crystal/label.cgi?id=136). More detail on the roll out and associated communications can be found on the UK Biobank website (http://biobank.ctsu.ox.ac.uk/crystal/refer.cgi?id=22).

### Defining Cases from the MHQ

Case definitions for the evaluation of the responses on the MHQ are detailed in Appendix 2 in supplementary materials. They arose either from the instruments used in the MHQ or by consensus criteria agreed by the working committee who wrote the MHQ. Diagnostic criteria were evaluated for depression (major depressive disorder), hypomania or mania, generalised anxiety disorder (GAD), alcohol use disorder, and post-traumatic stress disorder (PTSD). Addiction to substances and/or behaviour was defined based on self-report alone. Unusual experiences (describing potential symptoms of psychosis) and self-harm were also defined as phenomena that are important for phenotyping, but not disorder-specific. We combined outcomes to divide the cohort into five mood disorder groups, as shown in supplementary material Figure MD1.

Fulfilling the diagnostic criteria based on a self-report questionnaire does not allow us to rule out other psychiatric disorders, psychological or situational factors that might better explain the symptoms, and may have been elicited in a clinical evaluation. Therefore, we would regard any case classification arising from the MHQ as “likely”, rather than confirmed psychiatric disorder. The issue becomes particularly problematic for disorders that are less common in the population, such as bipolar affective disorder, where literature shows that using questionnaires to screen the population may over-estimate prevalence.^18^ Therefore, although we report the presence of hypomania/mania symptoms for the whole population, we only make the likely diagnosis of bipolar affective disorder in people with a history of depression, a sub-population where the prevalence of bipolar affective disorder is higher, and therefore screening questionnaires have better positive predictive values.^19^

### Analysis and Data Sharing

Data were supplied by UK Biobank on 8^th^ August 2017 under application number 16577. This data is open-access subject to the usual access procedures (www.ukbiobank.ac.uk).

Formal operational criteria (Appendix 2) written by KD based on consensus within the consortium (see Defining Cases above), with checks by JC, GB and MH (whole) and CD, NG, WL and DS (part). R code for analysis was developed by JC, with the code posted for comments during development, trialled on pilot data, and checked by KD and GB (whole). Portions of the data were analysed independently in parallel by other groups and subsequently compared (e.g. for mood disorders, by NG/BC). The code is freely available from Mendeley Data for the purpose of reproducing these analyses or developing further https://data.mendeley.com/datasets/kv677c2th4/3 (doi:10.17632/kv677c2th4.3).

We used R version 3.4.0-3.5.1 and MS Excel for analyses. We report numbers and proportions within the sample, and do not attempt to give population prevalence estimates, and 95% confidence intervals are therefore not shown.

## Results

The setting, recruitment and methods of selection of participants in UK Biobank have been published elsewhere.^1, 9^ For the mental health questionnaire study, 339,092 participants were sent an email invitation, and 157,366 (46% of those emailed) fully completed the questionnaire by July 2017 (available in August 2017) – which means that the MHQ had 31% coverage of the UK Biobank cohort. Figure 1 shows the flow chart of UK Biobank participants who completed the mental health questionnaire (MHQ). The median time for completion was 14 minutes, and 82% of respondents completed the questionnaire in under 25 minutes.

**Figure 1:**
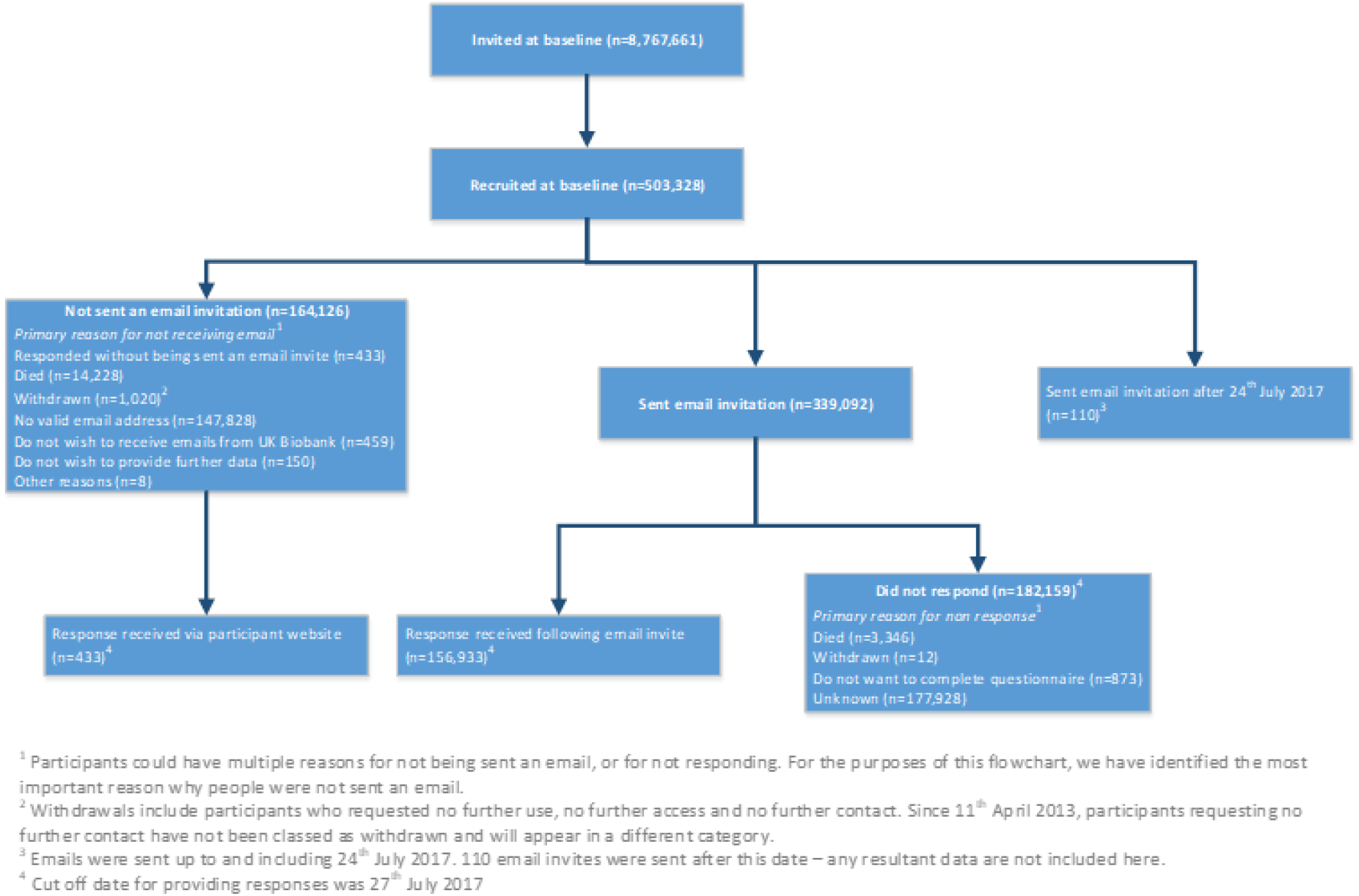
Flowchart of UK Biobank (UKB) participants from invitation to completion of mental health questionnaire (MHQ). Invitations were based on NHS registration, age and location. Numbers correct for July 2017.

Table SM2 in supplementary material shows participant characteristics for all UK Biobank participants and those who completed the MHQ compared to population-level data for UK residents in the same age range. They were different from the whole UK Biobank cohort and the general population by being better educated (e.g. 45% hold a degree vs 32% of UK Biobank participants vs 23% in the census), of higher socio-economic status according to job type, and healthier (longstanding illness or disability 28% vs 32% vs 37%), with lower rates of current smoking.

Table 1 shows that 34% of respondents reported they had received at least one psychiatric diagnosis from a professional at some time, and 12% had received two or more. The most commonly reported diagnosis was depression, followed by “anxiety or nerves”. Data are compared to the Health Survey for England (HSE) because this annual survey aims to report data that is a representative estimate for the population in England through its sample and weighting.^20^ The comparison shows that the pattern and prevalence of diagnosis is similar; for example, a depression diagnosis was self-reported by 21% of individuals in both samples, eating disorder by around 1%, and bipolar-related disorders by around 0.5%. The definition in the MHQ differed from that in the HSE for anxiety (MHQ definition was broader) and addiction (MHQ did not require professional diagnosis), and the higher overall prevalence in the UK Biobank MHQ compared to the HSE (35% vs 28%) may be due to those wider definitions.

**Table 1:** Respondent reports of mental health diagnoses by a professional (self-reported without physician diagnosis for addiction) compared to diagnoses reported in the Health Survey for England (HSE) 2014. NA = not reported.

|  | UK Biobank MHQ responses<br>(age 45-82y) |  | Health Survey for England (HSE)<br>(age 45-84y) |  |
| --- | --- | --- | --- | --- |
|  | N=157,366 | % in sample | N=3,272 | Prevalence (95% CI) |
| All psychotic disorders | 723 | 0.5 | 11 | 0.3 (0.2-0.6) |
| • *Schizophrenia* | 157 | 0.1 | NA |  |
| • *Any other type of psychosis or psychotic illness* | 604 | 0.4 | NA |  |
| Depression | 33424 | 21.2 | 679 | 20.8 (19.4-22.2) |
| Mania, hypomania, bipolar or manic-depression | 837 | 0.5 | 13 | 0.4 (0.2-0.7) |
| Anxiety, nerves or generalized anxiety disorder<sup>a</sup> | 22036 | 14.0 | 170 | 5.2 (4.5-6) |
| Panic attacks | 8704 | 5.5 | 262 | 8.0 (7.1-9.0) |
| Agoraphobia | 599 | 0.4 | NA |  |
| Social anxiety or social phobia | 1962 | 1.2 | NA |  |
| Any other phobia (eg disabling fear of heights or spiders) | 2153 | 1.4 | 27 | 0.8 (0.6-1.2) |
| Obsessive compulsive disorder (OCD) | 982 | 0.6 | 11 | 0.3 (0.2-0.6) |
| A personality disorder | 385 | 0.2 | 13 | 0.4 (0.2-0.7) |
| All eating disorders | 1851 | 1.2 | 26 | 0.8 (0.5-1.2) |
| • *Anorexia nervosa* | 891 | 0.6 | NA |  |
| • *Bulimia nervosa* | 503 | 0.3 | NA |  |
| • *Psychological over-eating or binge-eating* | 707 | 0.4 | NA |  |
| Autism, Asperger's or autistic spectrum disorder | 223 | 0.1 | NA |  |
| Attention deficit or attention deficit and hyperactivity disorder (ADD/ADHD) | 133 | 0.1 | 4 | 0.1 (0-0.3) |
| Any addiction or dependence | 9386 | 6.0 | NA |  |
| • Alcohol or drug addiction<sup>b</sup> | 5002 | 3.2 | 30 | 0.9 (0.6-1.3) |
| ○ *Physical alcohol dependence* | 946 | 0.6 | NA |  |
| *Summary* |  |  |  |  |
| None of above | 103346 | 65.7 | 2,356 | 72.0 (70.4-73.5) |
| 1+ above | 54020 | 34.3 | 916 | 28.0 (26.5-29.6) |
| 2+ above | 19400 | 12.3 | NA |  |

UK Biobank participants asked: “Have you been diagnosed with one or more of the following mental health problems by a professional, even if you don’t have it currently? (tick all that apply): By professional we mean: any doctor, nurse or person with specialist training (such as a psychologist or therapist). Please include disorders even if you did not need treatment for them or if you did not agree with the diagnosis.”
HSE participants asked to identify all the mental health conditions they had experienced, then asked whether they had been told by a doctor, psychiatrist or professional that they had it.
a) HSE participants were asked about Generalised Anxiety Disorder, and not about anxiety and nerves more generically
b) UK Biobank participants asked: “Have you been addicted to or dependent on one or more things, including substances (not cigarettes/coffee) or behaviours (such as gambling)?”. HSE definition of addiction includes physician diagnosis.

Table 2 shows that 45% of participants met criteria for one or more operationally defined syndromes. The most common was lifetime depression at 24%, followed by alcohol use disorder (21%), then generalised anxiety disorder (7%), PTSD (6%), unusual experiences (5%) and self-harm (4%). Hypomania/mania was the least common, at 2% of respondents. Table SM3 in supplementary material shows that women and men were approximately equally likely to have a history of one or more of the defined syndromes (women 44% vs men 46%), but differed as to which criteria were met: women were more likely to have a history of depression or anxiety disorder, while men were more likely meet criteria for a current alcohol use disorder (women 14% vs men 30%). Table 2 also shows the substantial comorbidity of defined syndromes. Notably, around three-quarters of participants who met criteria for lifetime anxiety disorder also met criteria for lifetime depression. Also, while individuals meeting criteria for PTSD had more than a two-fold risk of all of the lifetime syndromes compared to average, those identified with alcohol use disorder had little extra risk of lifetime syndromes.

**Table 2:** **Comorbidity** between operationally defined syndromes. Percentages refer to the proportion of participants with the row syndrome who also have column syndrome. See lettered table notes, and Appendix 2 for case definitions.

|  |  | Overall Prevalence | Comorbidity |  |  |  |  |  |  |
| --- | --- | --- | --- | --- | --- | --- | --- | --- | --- |
|  |  |  | Depression<sup>a</sup> | Hypomania / mania<sup>b</sup> | Anxiety disorder<sup>c</sup> | Unusual experiences<sup>d</sup> | Self-harm<sup>e</sup> | Alcohol use disorder<sup>f</sup> | PTSD<sup>g</sup> |
|  | **Total=** | 70892 (45%) | 37434 ( 24%) | 2396 ( 2%) | 11111 (7%) | 7803 (5%) | 6872 (4%) | 32602 (21%) | 10064 (6%) |
| **Lifetime history** | Depression<sup>a</sup> | 37434 ( 24%) |  | 1550 ( 4%) | 8444 ( 23%) | 3649 ( 10%) | 4240 ( 11%) | 8156 (22%) | 6373 ( 17%) |
|  | Hypomania/mania<sup>b</sup> | 2396 ( 2%) | 1550 ( 65%) |  | 778 ( 32%) | 598 ( 25%) | 453 ( 19%) | 660 (28%) | 657 ( 27%) |
|  | Anxiety disorder<sup>c</sup> | 11111 (7%) | 8444 (76%) | 778 (7%) |  | 1551 (14%) | 1704 (15%) | 2634 (24%) | 3274 (29%) |
|  | Unusual experiences<sup>d</sup> | 7803 (5%) | 3649 (47%) | 598 (8%) | 1551 (20%) |  | 1225 (16%) | 1784 (23%) | 1594 (20%) |
|  | Self-harm<sup>e</sup> | 6872 (4%) | 4240 (62%) | 453 (7%) | 1704 (25%) | 1225 (18%) |  | 1890 (28%) | 1719 (25%) |
| **Current** | Alcohol use disorder<sup>f</sup> | 32602 (21%) | 8156 (25%) | 660 (2%) | 2634 (8%) | 1784 (5%) | 1890 (6%) |  | 2572 (8%) |
|  | PTSD<sup>g</sup> | 10064 (6%) | 6373 (63%) | 657 (7%) | 3274 (33%) | 1594 (16%) | 1719 (17%) | 2572 (26%) |  |

a) Criteria met for major depressive disorder on CIDI-SF lifetime
b) Criteria met for hypomania / mania lasting for at least one week
c) Criteria met for generalised anxiety disorder on CIDI-SF lifetime
d) Reported potential hallucination or delusion at any point in their life
e) Reported deliberate self-harm at some point in their life, asked to report self-harm “whether or not you meant to end your life”
f) Score above cut-off (8+) on AUDIT during the last year
g) Criteria met for post-traumatic stress disorder on PCL-6 in the last month

In table 3, people meeting criteria for the lifetime occurrence of at least one of depression, bipolar disorder, generalised anxiety disorder, unusual experience or self-reported addiction are seen to be more likely than those without to come from a younger age group, report adverse life events, and have met criteria for loneliness or social isolation. They are more likely to have smoked cigarettes and/or used cannabis, and to have had a “longstanding illness” at baseline (although the presence of a mental disorder may have been the illness to which the participants refer in some cases), but all groups were equally likely to be achieving recommended levels of physical activity. Markers of deprivation (area-level deprivation and rented housing) are raised in groups with a history of mental disorders, especially bipolar affective disorder and addictions.

**Table 3.** Selected personal characteristics, socio-economic factors, risk factors and health behaviours by status for likely lifetime occurrence of operationally defined syndromes (people may be included in more than one category).

|  |  | No “lifetime”<br>criteria met<sup>a</sup><br>N=108,752 | Depression<sup>b</sup><br>N=37,434 | Bipolar type 1<sup>c</sup><br>N=931 | Anxiety disorder<br>(GAD)<sup>b</sup><br>N=11,111 | Unusual<br>experiences<sup>d</sup><br>N=7,803 | Addiction<sup>e</sup><br>N=9,386 |
| --- | --- | --- | --- | --- | --- | --- | --- |
| Personal Characteristics |  |  |  |  |  |  |  |
| Age<sup>f</sup> | 45-54 | 14364 (13%) | 7145 (19%) | 228 (24%) | 2348 (21%) | 1485 (19%) | 2013 (21%) |
|  | 55-64 | 33307 (31%) | 14809 (40%) | 417 (45%) | 4470 (40%) | 2904 (37%) | 3428 (37%) |
|  | 65-74 | 51705 (48%) | 13739 (37%) | 261 (28%) | 3892 (35%) | 2960 (38%) | 3466 (37%) |
|  | 75+ (oldest is 82) | 9376 (9%) | 1741 (5%) | 25 (3%) | 401 (4%) | 454 (6%) | 479 (5%) |
| Gender | female | 57556 (53%) | 25815 (69%) | 532 (57%) | 7404 (67%) | 4718 (60%) | 4556 (49%) |
| Ethnicity | white | 105072 (97%) | 36297 (97%) | 892 (96%) | 10749 (97%) | 7503 (96%) | 9037 (96%) |
| Townsend Deprivation Score<sup>g</sup> | most deprived (TDS ≥ +2) | 11783 (11%) | 5656 (15%) | 201 (22%) | 1856 (17%) | 1426 (18%) | 1941 (21%) |
| Highest qualification | degree | 48700 (45%) | 16939 (45%) | 425 (46%) | 5071 (46%) | 3646 (47%) | 4531 (48%) |
| Housing tenure | rent<sup>h</sup> | 4162 (4%) | 2906 (8%) | 155 (17%) | 1026 (9%) | 854 (11%) | 1109 (12%) |
| Known risk factors |  |  |  |  |  |  |  |
| Neuroticism score<sup>i</sup> | mean (SD) | 3.2 (2.8) | 5.6 (3.3) | 3.8 (3.1) | 7.1 (3.3) | 5.2 (3.5) | 5.4 (3.5) |
| Adverse life experiences | childhood screen<sup>j</sup> | 43913 (40%) | 21144 (56%) | 638 (69%) | 6931 (62%) | 4783 (61%) | 5800 (62%) |
|  | adult screen<sup>k</sup> | 50226 (46%) | 23893 (64%) | 685 (74%) | 7581 (68%) | 5284 (71%) | 6303 (67%) |
|  | trauma exposure<sup>l</sup> | 50771 (47%) | 22166 (59%) | 665 (71%) | 6877 (62%) | 5439 (70%) | 6278 (67%) |
| Social connection | loneliness<sup>l,m</sup> | 2976 (3%) | 2367 (6%) | 94 (10%) | 971 (9%) | 570 (7%) | 669 (7%) |
|  | social isolation<sup>i</sup> | 7793 (7%) | 3827 (10%) | 126 (14%) | 1173 (11%) | 931 (12%) | 1200 (13%) |
| Illness | longstanding illness,<br>disability or infirmity<sup>j</sup> | 26341 (24%) | 13363 (36%) | 503 (54%) | 4581 (41%) | 3242 (42%) | 3588 (38%) |
| Health-behaviours |  |  |  |  |  |  |  |
| Smoking status<sup>i</sup> | current | 6235 (6%) | 3638 (10%) | 158 (17%) | 1194 (11%) | 837 (11%) | 1916 (20%) |
| Cannabis use (lifetime) | daily (ever) | 868 (1%) | 914 (2%) | 63 (7%) | 346 (3%) | 258 (3%) | 867 (9%) |
| Physical activity<sup>i</sup> | moderate activity ≥ three<br>times a week | 39677 (36%) | 13988 (37%) | 345 (37%) | 4174 (38%) | 2846 (36%) | 3602 (38%) |

a) Criteria not met for depression, GAD, unusual experiences or addiction.
b) Criteria met for disorder on CIDI-SF lifetime
c) Criteria met for both lifetime depression and lifetime mania
d) Reported potential hallucination or delusion at any point in their life
e) Positively endorsed: “Have you been addicted to or dependent on one or more things, including substances (not cigarettes/coffee) or behaviours (such as gambling)?”
f) Age when mental health questionnaire completed, derived from date of birth
g) Townsend material Deprivation Score is based on postcode areas
h) Includes rent social and rent private, excludes other categories of housing tenure
- i) From baseline assessment 2006-10 - j) Criteria met for possible abuse or neglect on Childhood Trauma Screener - k) Criteria met for adverse situations as an adult: lack of confiding relationship, abusive relationships and money problems - l) Reports one or more of six situations that are known to be triggers for trauma-related disorders - m) There is some overlap between the adult screen and loneliness screen, which both ask about confiding relationships: adult screen includes lack of confiding relationship over the adult lifetime; loneliness includes lack of confiding relationship at the time of baseline assessment

The supplementary materials have a section on mood disorder, showing the results of analyses of MHQ participants by likely disorder categories (figure MD1). Table MD1 shows the features of these groups. The characteristics of people who meet diagnostic criteria for depression appear to be shared by those with subthreshold depressive symptoms. Table MD2 shows comorbidity, and demonstrates a gradient effect in the presence of a non-depression syndrome rising from 22% in no depression (mainly alcohol use disorder) to 50% in recurrent depression. Table MD3 shows that people with a history of depression or bipolar affective disorder tend to have worse scores for current mental health.

## Discussion

This paper has described the development, implementation and principal descriptive findings from the UK Biobank Mental Health Questionnaire (MHQ). The implementation of this questionnaire demonstrates that a web-based questionnaire is an acceptable means of collecting mental health information at low cost and large scale. Whilst the data collection methods might force more limited data acquisition than conventional interview methods, with associated uncertainties in true diagnostic categorisation, we suggest that the survey achieved an acceptable trade-off between depth of phenotypic information and scale of sample size.

The MHQ achieved a participation rate of 31% of the original UK Biobank participants and 46% of those emailed. This response rate is substantially higher than previous UK Biobank questionnaires, largely owing to the attention paid to ensure the acceptability of the invitation and questionnaire and the efficient use of reminders. Those who completed the MHQ appear to be better educated and have higher socio-economic status (on all measures: job title, household income, home ownership and area level deprivation) than those recruited into UK Biobank overall, and the UK population. Despite this, we found that rates of self-report diagnoses were similar to population estimates from the Health Survey England. The patterns of association between disorders and demographics were also broadly as predicted by previous research, which adds to the face validity of the questionnaire. For example, depression and anxiety were more common in women, while addiction and alcohol misuse were more common in men, and all disorders were less common in respondents older than 65 years. The decrease in prevalence of lifetime disorder with increasing age has been previously noted in cross-sectional studies, although the causes and implications are not clearly understood.^21, 22^ The high level of alcohol use disorder found using the AUDIT (Alcohol Use Disorder Identification Tool) is consistent with the Adult Psychiatric Morbidity Survey 2014, where they comment on increasing numbers in older age groups since 2007.^23^

The ‘healthy volunteer’ selection bias within the UK Biobank has been previously explored^24^. The impact of selection biases on disease prevalence are likely to be particularly strong for mental disorders, where disorder status or symptoms may influence participation in research,^25, 26^ since many risk factors for these disorders, including genetic risk as polygenic risk score for mental disorder can be associated with non-participation.^27^ Therefore, the results of the MHQ should not be used to provide prevalence estimates. However, the pattern of the measured risk factors among the participants with mental disorders in the MHQ, including neuroticism, trauma, loneliness and housing tenure, was in accordance with established literature, supporting the use of the data to study the relationships between exposures and outcomes. Previous work on health surveys with selection bias due to non-participation, including UK Biobank, have indicated that they can be used to give estimates of association,^11, 25, 28^ although bias may occur in some cases.^29, 30^ For example, the relative underparticipation of unskilled workers in the MHQ (around one-fifth of the proportion in the population) could mask an association with a variable that was related to unskilled work.

## Strengths and Limitations

We developed a questionnaire through a consensus approach with clear aims of capturing enough data to characterise participants as having a lifetime history of depression and other phenotypes. Validated instruments were used where possible. The consortium working on the questionnaire included mental health researchers and members of the UK Biobank team working in collaboration to develop the optimum approach. The derived variables of likely categorical diagnoses will be added to the UK Biobank resource, facilitating those less familiar with mental health to use the results efficiently. The UK Biobank data, including that from the MHQ, is available to researchers and we have made the code used to derive the results in this paper freely available, allowing other researchers both to both query our findings and build upon them for their own work.

The ‘healthy volunteer’ effect may limit applications of the data. Due to restrictions of time and space, the questionnaire was limited in the topics it could cover. The focus of the questionnaire was on categorical diagnoses rather than dimensional traits, which will tend to confirm conventional ICD/DSM nosology of psychiatric disorder and may not suit some research^31^. In particular, tools were chosen that are based on DSM-IV disorders, which reflects current practice (for example National Institute for Health and Care Excellence (NICE) guidelines on depression and anxiety use DSM IV definitions)^32, 33^. Of the disorders with operational classification, all would generalise to DSM-5, except post-traumatic stress disorder^34^, and the concepts are valid for ICD-10 disorders, although the threshold of disorder may be different, e.g. depression is diagnosed with fewer symptoms in DSM than ICD^32^. The questionnaire was heavily reliant on participant report, which may be affected by stigma of reporting psychiatric symptoms, and tends to underestimate lifetime prevalence through forgetting or re-evaluating distant events^11, 21, 35^. We hope that some of these shortcomings can be addressed in the future by a more fine-grained analyses of the MHQ data, supplemented with other data from UK Biobank to create a richer picture of mental health in the cohort.

## Conclusions

UK Biobank offers a unique opportunity to research common disorders in a well characterised longitudinal cohort of UK adults. A detailed mental health questionnaire has now been completed by 157,366 participants, including self-report, operationally defined lifetime disorder status, and detailed phenotype information on mood disorder. The proportion of cases and the patterns of participants experiencing symptoms and disorders was as expected despite a “healthy volunteer” selection bias. Further work on mental health phenotyping for UK Biobank includes validation of Hospital Episode Statistics for mental health diagnoses,^12^ incorporation of general practice records, triangulation of health record and questionnaire data^36^, and investigation of further putative phenotypes^37^. Existing projects utilising UK Biobank mental health data can be seen in a searchable database of approved research (http://www.ukbiobank.ac.uk/approved-research/).

This study also demonstrates of the substantial burden of mental disorders, including potentially dangerous patterns of alcohol consumption. Given the known impact of mental health on physical health, mental health data and its associations should interest researchers from every biomedical specialty. This study suggests that UK Biobank could be a powerful tool for such studies, and since it is open to all *bona fide* health researchers for work in the public good, we hope this study will inspire both existing and new users of UK Biobank.

## Data Availability

This data is open-access subject to the usual UK Biobank access procedures (www.ukbiobank.ac.uk). The code is freely available from Mendeley Data.

https://data.mendeley.com/datasets/kv677c2th4/3

## Acknowledgements

We thank the staff and participants of UK Biobank, the PROTECT study and the SLaM Service Users Group for their participation.

## Author Contribution

All authors’ contribution met the ICMJE criteria for authorship. KD, GB, EF, LH, AJ, RM, AM, DS, CS, SZ and MH designed the study. NA and JH co-ordinated the delivery of the questionnaire. GB and JC curated data. KD, JC, MA, BC, CD, NG, WL, RP, DS and JW contributed to analysis. KD, JC and MH wrote the paper. All authors critically edited. All authors agreed to submission.

## Data Availability

Available from UK Biobank subject to standard procedures (www.ukbiobank.ac.uk). Code for replication available from Mendeley Data (doi:10.17632/kv677c2th4.3).

